# Joint hypermobility and its relevance to common mental illness in adolescents: a population-based longitudinal study

**DOI:** 10.1101/2020.09.14.20191130

**Authors:** Jessica A Eccles, Hannah E Scott, Kevin A Davies, Rod Bond, Anthony S David, Neil A. Harrison, Hugo D Critchley

## Abstract

**Importance:** Depression and anxiety are common mood disorders that show higher prevalence in adults with joint hypermobility, a consequence of a constitutional variant of connective tissue structure. In adolescents, an association between mood disorder and hypermobility may enhance the potential understanding of risk factors for emotional disorder and provide opportunities for early intervention approaches.

**Objective:** To test the hypothesis that joint hypermobility, a consequence of constitutional variant of connective tissue, is associated with common mental illness in adolescents.

**Design:** This was a longitudinal prospective study.

**Setting:** The Avon Longitudinal Study of Parents and Children (ALSPAC) is a prospective ongoing general population birth cohort study based in Avon County, England.

**Participants:** The original data set comprised 6105 individuals from the cohort with data available on joint hypermobility at age 14 years; a sub-sample (n=3803) had later psychiatric assessments.

**Measurement of Exposure:** Joint hypermobility was measured by physical examination at age 14 and 18 years, using the Beighton Scale

**Main Outcome and Measures:** Participants were assessed at age 18 years. ICD-10 diagnoses of Depression and Anxiety were obtained using the Clinical Interview Schedule-Revised (CIS-R) and levels of anxiety quantified using the Anxiety Sensitivity Index (ASI)

**Results:** Presence of generalized joint hypermobility (GJH) at age 14 years predicted depression at 18 years in males (Odds Ratio (OR) 2.10; 95%CI, 1.17-3.76) but not females After accounting for missing data it was determined that this relationship was mediated by heart rate, a potential measure of physiological arousal. Symptomatic hypermobility ((GJH plus chronic widespread pain (CWP)) at age 18 years was further associated with the presence of anxiety disorder (OR 3.13; 95% CI 1.52-6.46) and level of anxiety (Beta = 0.056, t(3315)=3.27), depressive disorder (adjusted OR 3.52; 95%CI, 1.67 – 7.40) and degree of psychiatric symptomatology (Beta 0.096, t(2064)=4.38)

**Conclusions and relevance:** Generalized joint hypermobility and symptomatic hypermobility are associated with common mental disorders in adolescence. Consideration of hypermobility may provide important opportunities for intervention to mitigate psychiatric disorder.

**KEY POINTS:** *Question:* Is joint hypermobility related to depression and anxiety in adolescence?

*Findings:* In this longitudinal study presence of generalized joint hypermobility at 14 predicted subsequent adolescent depression in males only and this relationship was mediated by heart rate. At age 18 symptomatic hypermobility was associated with both presence of anxiety and depression and psychiatric symptom severity.

*Meaning:* Generalized joint hypermobility and symptomatic hypermobility are associated with common mental disorders in adolescence.

## INTRODUCTION

Depression and anxiety are among the leading causes of morbidity worldwide ^1^, and typically first manifest during adolescence ^2^. Longitudinal studies provide insight into the development of such common disorders ^3-10^. Identification of risk factors is of vital importance for developing preventative and early intervention strategies to mitigate future morbidity and promote well-being ^11,12^.

Joint hypermobility ^13^ occurs as a consequence of a frequent variant of connective tissue, usually manifesting as an extended range of motion in multiple joints, i.e. generalized joint laxity ^14^ or generalized joint hypermobility (GJH)^15^, typically assessed using the Beighton scale ^16^. Hypermobility decreases with age, and is more common in females ^17^. General population estimates approximately 20% of adolescents ^14^ and adults ^18^ are hypermobile. Clinically, hypermobility also occurs in heritable connective tissue disorders (HDCT), including Ehlers-Danlos Syndrome (EDS), Marfan’s Syndrome and Osteogenesis Imperfecta ^19,20^. A diagnosis of symptomatic hypermobility can be made when hypermobility is associated with significant musculoskeletal or connective tissue symptoms. This classification arises from consensus changes to terminology and nomenclature ^21^, encompassing Hypermobile EDS (hEDS) ^22^ (formerly known as EDS hypermobility type/EDS type-III/Joint Hypermobility Syndrome (JHS) ^23,24^) and Hypermobility Spectrum Disorder^21^. Extra-articular manifestations of hypermobility are increasingly recognized ^20^, including psychiatric disorder ^25,26^.

Hypermobility appears to bestow constitutional vulnerability to psychiatric conditions ^27,28^. Interestingly, this has now been described in the domestic dog ^29^, wherein hypermobility is linked to emotional arousal. In human adults, individuals with hypermobility are over-represented in common mental disorders; panic, anxiety and depression (OR 6.72, 4.39 and 4.10) ^30^. A population-wide study in adults demonstrated significantly greater risk of depression (RR 3.4) in individuals with EDS/JHS ^31^. Moreover, hypermobility is associated with structural and functional differences in brain regions implicated in physiological and experiential aspects of emotion, notably amygdala ^32^ and insula ^33,34^. Dysautonomia is also observed in hypermobility^35,36^, typically, as ‘postural tachycardia syndrome’ which has phenomenological overlap with anxiety ^37^. Hypermobility is a risk factor for chronic widespread pain ^38^ and is commonly expressed in neurodevelopmental disorders (e.g. ^17,39-43^). Nevertheless, only a limited literature explores the link between hypermobility and other common psychological conditions in children and adolescents.

The objective of this study was to use a birth cohort to test longitudinally for an association between hypermobility in early adolescence and subsequent risk of anxiety and depression in later adolescence.

## METHODS

### Description of the cohort

The Avon Longitudinal Study of Parents and Children (ALSPAC), birth cohort consists of 14062 live births from women living in Avon County, a geographically defined region in southwest England, with expected dates of delivery between April 1991 and December 1992 (http://www.bristol.ac.uk/alspac) ^44,45^. Parents completed regular postal questionnaires about all aspects of their child’s health and development from birth. From the age of 7 years, children attended annual assessment clinics during which they participated in a variety of physical tests and face-to-face interviews, receiving ethics approval from the ALSPAC Law and Ethics Committee. All participants provided written informed consent. All data is available on the ALSPAC data dictionary and variable search tool (http://www.bristol.ac.uk/alspac/researchers/our-data/)

Analyses for this study are based on two samples – one for a predictor model, and the second for an association model. The risk set for the predictor model included the 6105 adolescents who underwent a complete hypermobility assessment in the clinic as 14-year old adolescents. Of this risk set, 5731 adolescents also had heart rate data recorded at the same time-point. Of these, 3803 went on to complete assessments for depression and anxiety (CIS-R) aged 18 years. Our principle longitudinal analyses were based on this later sample and were repeated after using maximum likelihood estimation to account for missing data. The second (association) sample included the 4390 adolescents who completed assessments for both hypermobility and the CIS-R aged 18 years. Finally, a subset of 3557 adolescents from the association sample who also completed assessment for chronic widespread pain at the same 18-year time point were additionally analyzed.

### Assessment and definition of hypermobility (*GJH*)

Generalized joint hypermobility was measured in a dedicated research clinic by trained assessors using the modified Beighton 9-point scoring system ^16^ at two time points: age 14 and age 18. Each joint was assessed separately. The fifth metacarpophalangeal joint was scored as hypermobile if it could be extended >90°, the thumb if it could be opposed to the wrist, and the elbows and knees if they could be extended >10°. The spine was scored as hypermobile if both palms could be placed flat on the floor with the knees straight. Scores were recorded for individual joints, and a total score (maximum of 9) was ascertained. In common with other investigators we used a cut-off of ≥4 hypermobile joints as a dichotomised definition of generalized joint hypermobility ^14,46-48^. Heart rate was measured in the same assessment clinics.The only formal research validated diagnosis of symptomatic joint hypermobility, e.g. joint hypermobility syndrome (JHS) are based on the revised Brighton criteria ^24^, requiring a combination of major and minor criteria consisting of generalized joint hypermobility and significant musculoskeletal or connective tissue symptoms. Typically, JHS was diagnosed in the presence two major criteria (one being a Beighton score of 4 or more). However, diagnosis could also be made on the basis of one major and two minor criteria, or alternately four minor criteria. Not all minor criteria were recorded in the cohort. We therefore constructed a narrow definition of ‘likely JHS’ based on the presence of generalized joint hypermobility and concomitant chronic widespread pain (CWP)^49^ at age 18 years. This was operationalized as meeting the two major criteria of the Brighton JHS assessment ^24^.

### Assessment of psychiatric conditions

Depression and anxiety were measured at age 18 using the self-adminstered computerized version of the Clinical Interview Schedule-Revised (CIS-R). CIS-R is a widely used standardized tool for measuring depression and anxiety in community samples ^50^, including this cohort ^7,9,10^. The CIS-R scores symptoms to assign International Classification of Diseases-10 (ICD-10) diagnoses of depression and anxiety disorders ^51,52^. We derived binary variables indicating: 1) presence or absence of a primary diagnosis of major depression and 2) presence or absence of a primary diagnosis of any anxiety disorder (including generalized anxiety disorder, panic disorder, agoraphobia, social phobia and specific phobia). These variables were assessed independently. In addition, we derived two severity indices: 3) degree of psychiatric symptomatology from the total CIS-R score, and 4) degree of anxiety from the Anxiety Sensitivity Index (total of mental and physical concerns subscale).

### Statistical Analysis

Statistical analyses were conducted in IBM SPSS Statistics Version 24 and Mplus Version 8^53^.

To test the hypothesis that hypermobility was a predictor for subsequent anxiety or depression at age 18 years we firstly we used separate logistic regression models to calculate the odds ratios (ORs) and 95% confidence intervals (CI) for depressive disorder and/or anxiety disorder at age 18 years for individuals with and without generalized joint hypermobility at 14 years. As hypermobility is known to be more common in females, analyses for males and females were performed separately. Separate logistic regression models were used to test the effect of heart rate at age 14 years on both generalized joint hypermobility at aged 14 years and on the later expression of depression and anxiety at age 18 yrs.Secondly, we performed a mediation analysis ^54^ with psychiatric disorder at age 18 years as the outcome variable, generalized joint hypermobility at age 14 years as the predictor, and heart rate at age 14 years as a mediating variable. Probit regression was used as the outcome variable was categorical and parameter estimates computed from maximum likelihood estimation to account for missing data. Bootstrapping (n=2000) was applied with the generation of 95% bias-corrected bootstrap confidence intervals to test inferentially for direct and indirect mediation effects. Analyses were performed separately for males and females. Mediation analyses were controlled for mean-centered age at assessment and body mass index, as both were associated with the potential mediator (heart rate).

In order to test for association between symptomatic hypermobility (likely JHS) and concurrent psychiatric disorder separate logistic regression models were used to calculate ORs and 95% CI for depression and anxiety disorder at age 18 years in people with and without symptomatic joint hypermobility (likely JHS). Linear regression tested for differences in anxiety sensitivity and total CIS-R score between individuals with and without symptomatic joint hypermobility. As data was collected at the same time point, these analyses were not subject to maximum likelihood estimation.

## RESULTS

### Participant characteristics

Of the 6105 individuals assessed for hypermobility at 14 years, 3803 underwent assessment for depression and anxiety at 18 yrs.and 3557 had assessments for chronic widespread pain and hypermobility. See Table 1 for a description of the cohort

**Table 1:** Characteristics of the sample

| Timepoint | Evaluation | n | Age (mean±s.d, range) | % male |
| --- | --- | --- | --- | --- |
| **Risk sample** |  |  |  |  |
| Clinic aged 14 | Generalized joint hypermobility<br>Heart rate | 6105 | 13.8±0.2 yrs, 12.5-15.2 | 49 |
| Clinic aged 18 | Depression<br>Anxiety | 3803 | 17.7±0.4 yrs, 16.3-19.3 | 45 |
| **Association sample** |  |  |  |  |
| Clinic aged 18 | Likely JHS<br>Depression<br>Anxiety | 3557 | 17.7±0.4 yrs, 16.3-19.3 | 42 |

### Prevalence of hypermobility and association with age and sex

Of those assessed at 14 years, 1175 (19%) had generalized joint hypermobility as reported elsewhere ^14^. This was more common in females (n=856, 28%) compared to males (n=319, 11%), (OR 3.20, 95% CI 2.78-3.68, p=<0.001) and not associated with variation in age.

Of those who had assessments for hypermobility and chronic widespread pain at 18 years, 36 (1%) had ‘likely JHS’. This was significantly more common in females; only 3 were male (OR 8.04, 95% CI 2.46 −26.25, p=0.001) and was not associated with variation in age.

### Risk set analysis

#### Risk of anxiety and depression at age 18 according to hypermobility status at age 14

Of those assessed for hypermobility at 14 years, a sub-goup underwent assessment for depression and anxiety at 18 yrs (see Table 1). Of these 3803 participants, 293 (7.7%) met ICD-10 diagnostic criteria for depression; 215 female. Age at assessment for was not associated with diagnosis.

Across the whole group, generalized joint hypermobility was a significant predictor of depression at 18 (OR 1.70 95% CI 1.31 – 2.22, p=0.001). Of the 293 individuals who met ICD-10 criteria for depression, 86 (29%) were hypermobile (95%CI 24.43 – 34.48%); of those not depressed (n=3510), significantly fewer, 688 (20%) were hypermobile (95% CI 18.31 – 20.96%). There was a significant interaction of generalized joint hypermobility and sex on depression (F_(2,3802)_ =3.73, p=0.024). This result was driven by a significant effect in males only (OR 2.10, 95%CI 1.17-3.76, p=0.013). (Figure 1; Table 2).

**Figure 1:**
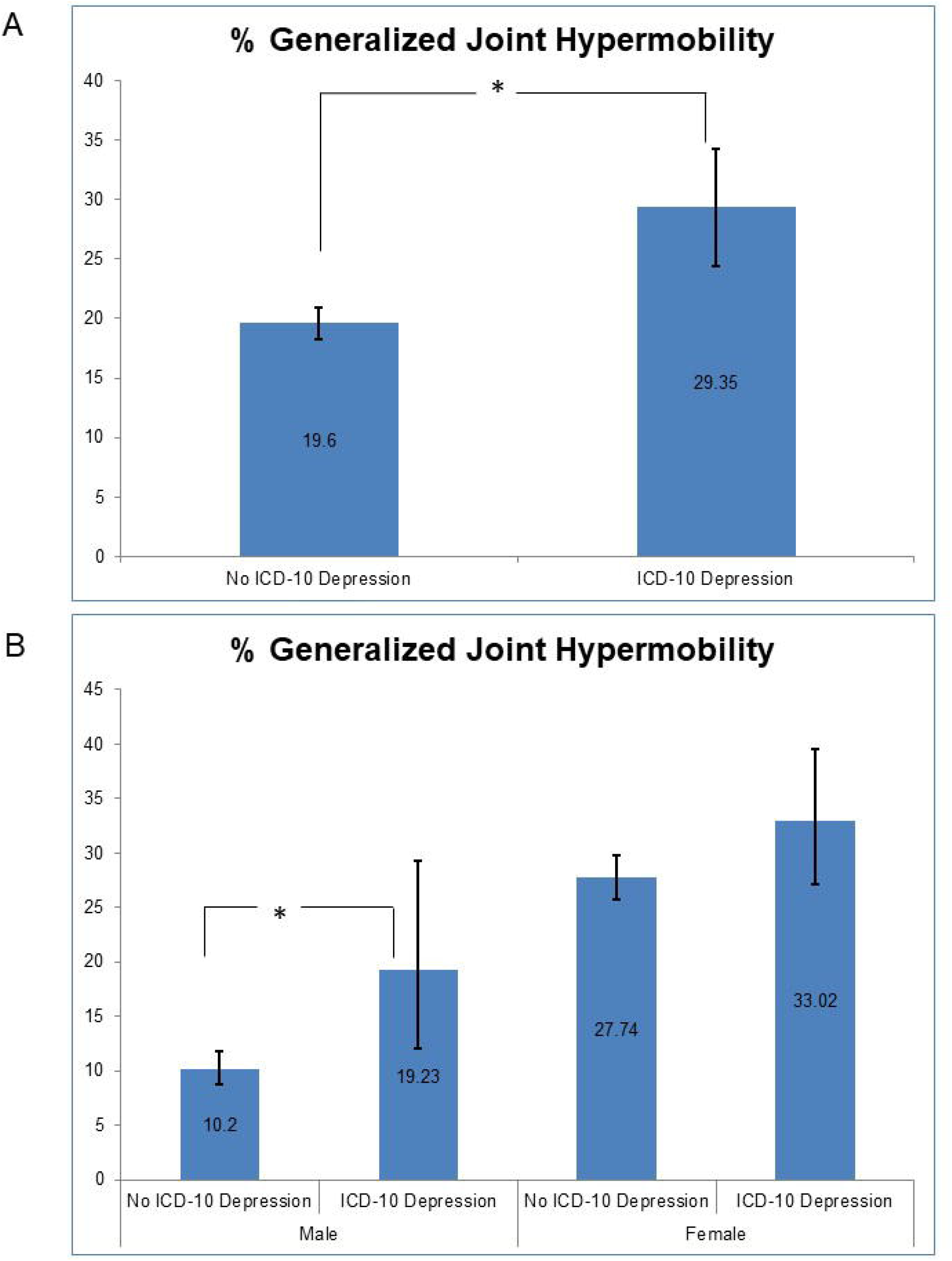
Relationship between generalized joint hypermobility and depression: Percentage of each group (ICD-10 Depression or not) who had generalized joint hypermobility, demonstrating significantly higher percentage of joint hypermobility in those who were depressed compared to those who were not across the whole group and in males only. Error bars represent 95% confidence intervals of percentage with generalized joint hypermobility; Mean values are indicated within bars.

Three hundred and sixty six adolescents (9.6% 95% CI 8.72 – 10.6) met ICD-10 diagnostic criteria for anxiety disorder; 265 female. Age was not associated with diagnosis. There was no association between anxiety disorder and hypermobility at 14 (Table 2).

**Table 2:**
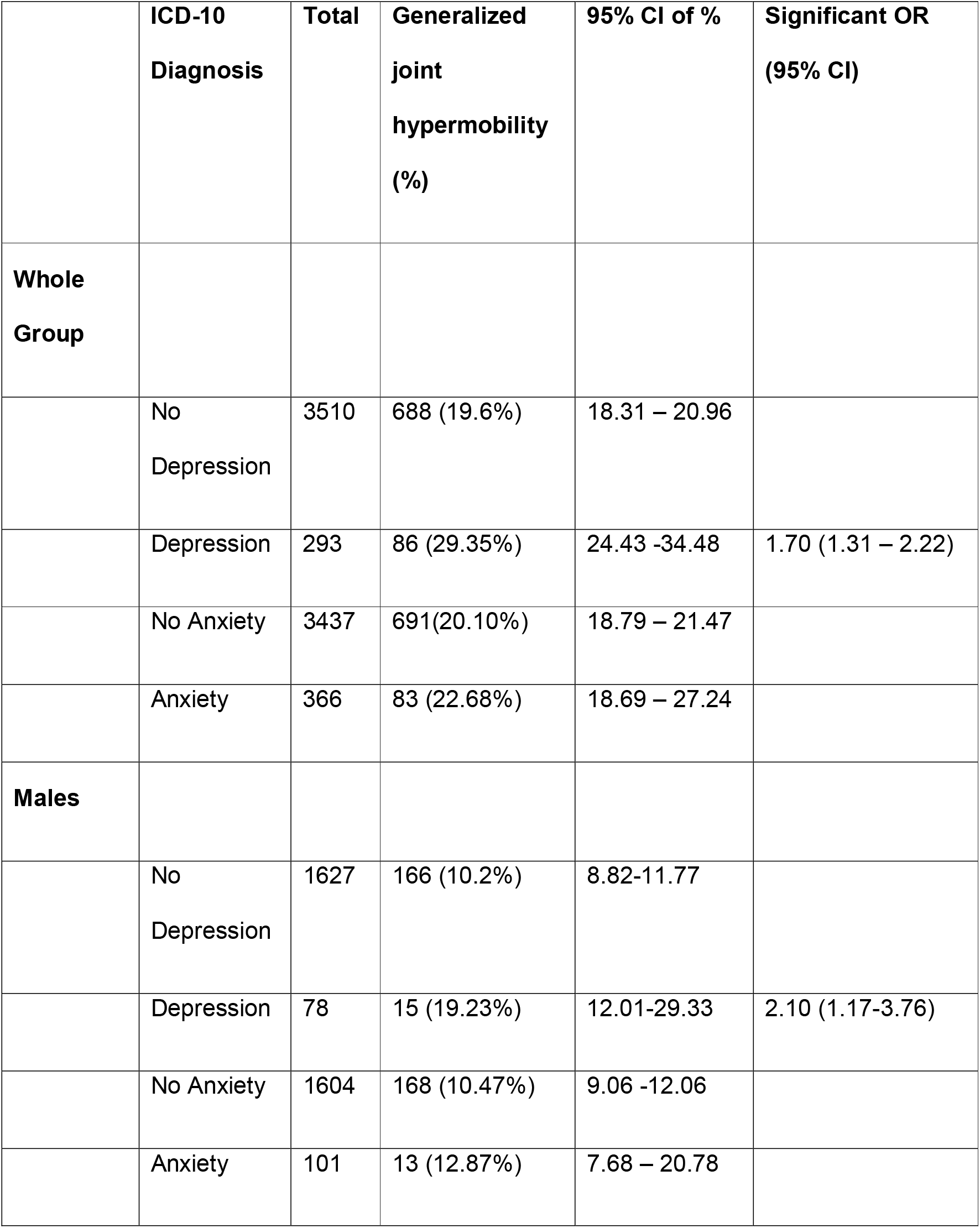
Relationship between generalized joint hypermobility at age 14 years and depression and anxiety aged 18 years, with significant odds ratio of psychiatric disorder given hypermobility. Relationship between symptomatic hypermobility and psychiatric disorder at age 18 with adjusted odds ratio of psychiatric disorder given hypermobility (adjusted for sex).

#### Associations with heart rate at age 14yrs

In this risk set assessed for hypermobility at 14 yrs, 5731(93.9%, 2832 male, also had heart rate measurements. Heart rate (mean bpm, sd) was significantly higher in females (83.92, 11.94) (78.6, 11.44, t(5725)=17.18, p=<0.001), and was significantly associated with age (r(5729)=-0.32, p=0.016) and BMI (r(5722)=0.081, p=<0.001) at assessment. After correcting for age and BMI, heart rate at 14 yrs was significantly associated with both hypermobility age 14 yrs (OR 1.01, 95%Ci 1.01 −1.02, p=<0.001) and also depression at 18 (0R=1.01, 95% CI 1.00-1.02, p =0.007).

#### Risk of depression at age 18 after accounting for missing data

Of the risk set at 14 yrs, 2305 (37.7%) cases were missing for assessment of subsequent depression at 18 yrs. After accounting for missing data, the OR of depression at 18 yrs in both males and females, given hypermobility at 14 yrs was 1.28 (b=0.246, 95% CI 1.12-1.47, p=<0.001). In males, the OR was 1.38 (b=0.32, 95% CI (0.97 – 1.79, p=0.036). There was no significant effect in females.

#### Mediation Analysis after accounting for missing data

There was evidence of mediation of the relationship between hypermobility and depression by heart rate in males only as the indirect effect odds ratio of hypermobility on depression is estimated as 1.050 and is significant because the 95% confidence interval does not cover one (1.011, 1.125). For full details see Table 3

**Table 3:**
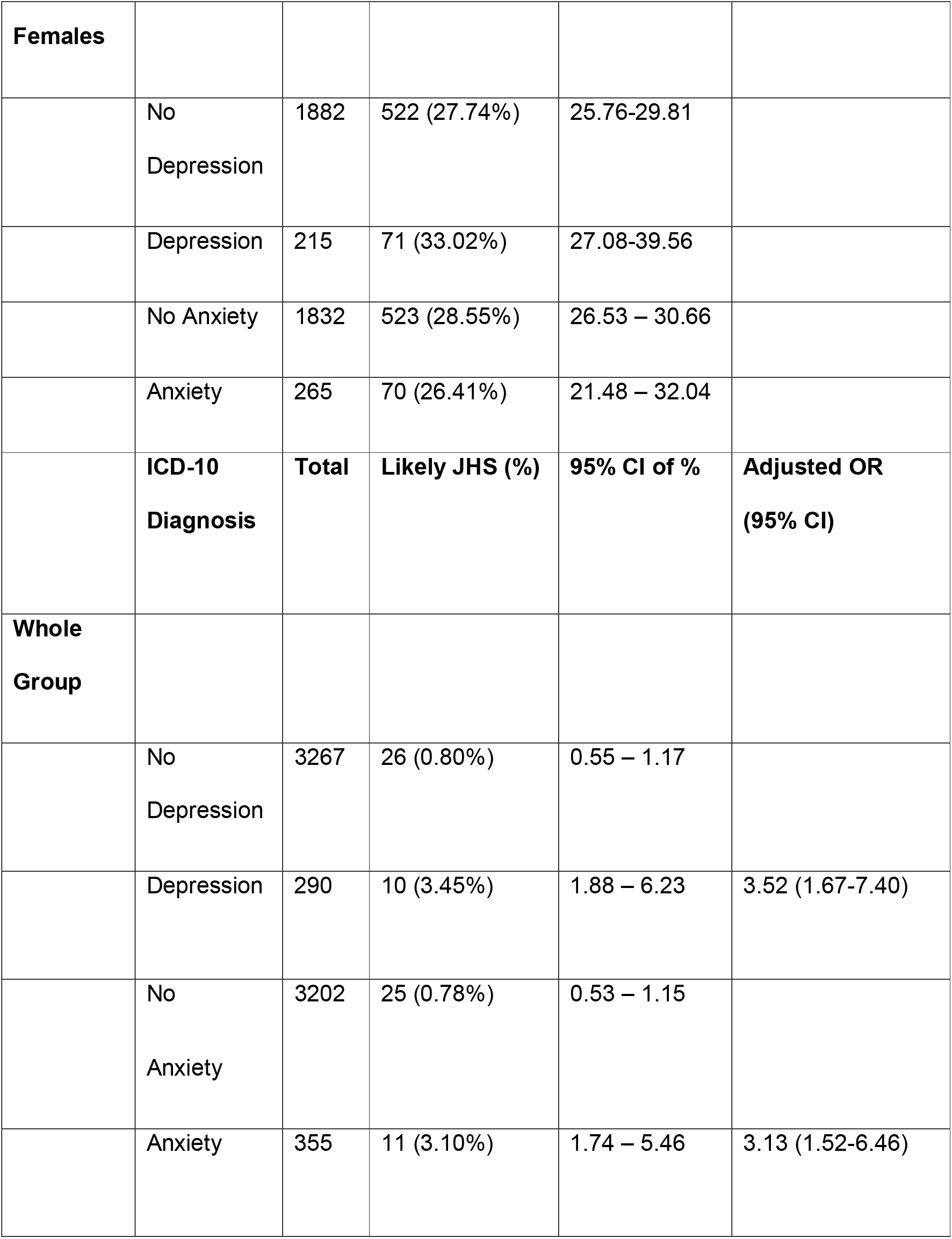
Bootstrap confidence intervals for hypermobility effects using probit regression for the outcome depression in males only with heart rate as potential mediator, demonstrating mediaton by heart rate.

#### Association set

##### Symptomatic hypermobility (likely JHS) and depression at age 18

Across the whole group ‘likely JHS’ was significantly associated with depression at 18 (OR 4.45 95% CI 2.13 – 9.33, p=<0.001). Of those (n=290) who met ICD-10 criteria for depression, 10 (3.45%) had ‘likely JHS’ (95%CI 1.88 – 6.23); of those not depressed (n=3267), a significantly lower proportion, 26 (0.80%) had ‘likely JHS’ (95% CI 0.55 −1.17). (Figure 2, Table 1)

**Figure 2:**
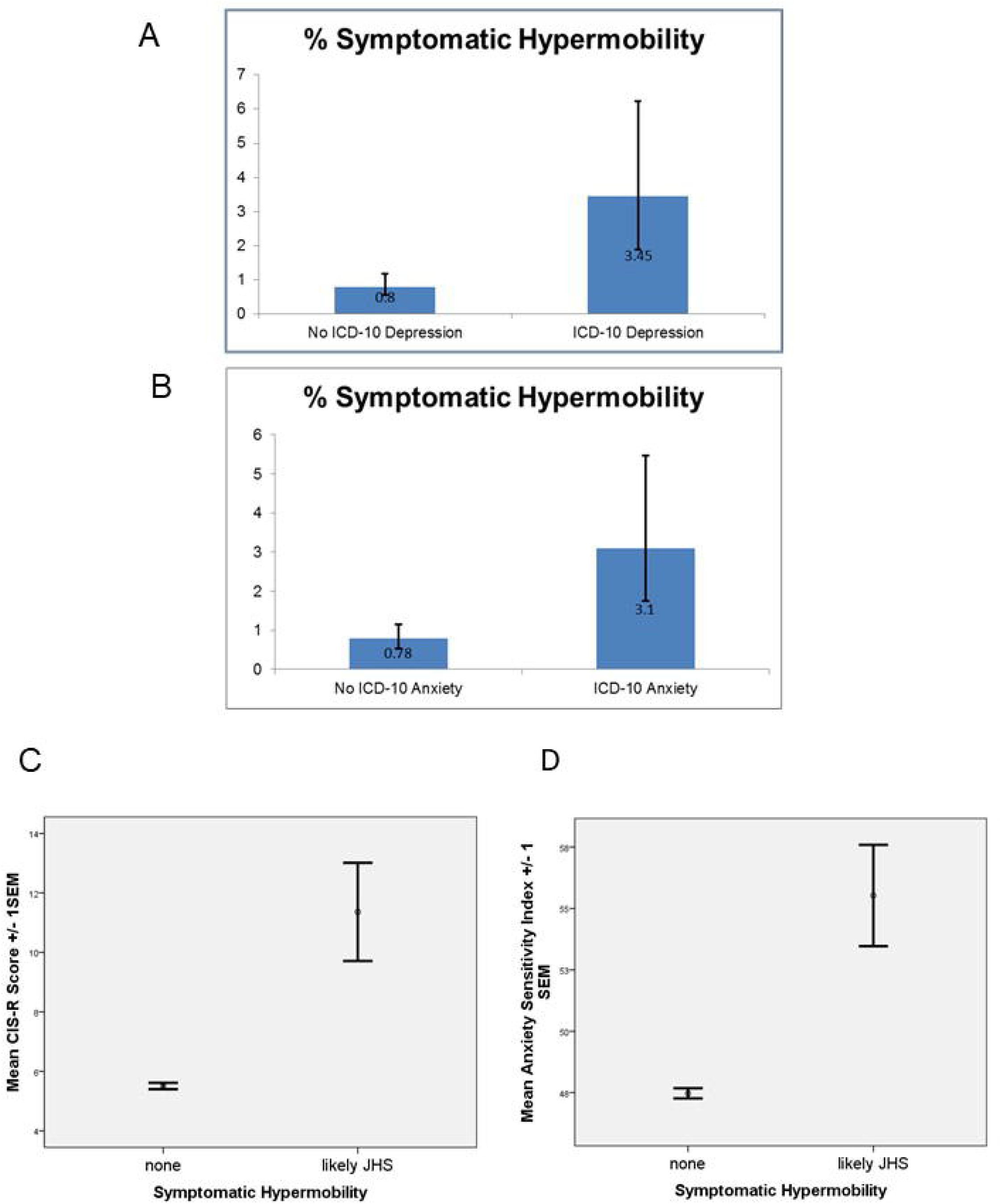
Significant association between symptomatic hypermobility and A) depressive disorder and B) anxiety disorder with significantly higher psychopathology scores C) and anxiety scores D) associated with symptomatic hypermobility at age 18.

In analyses corrected for sex, OR of depression at 18 given ‘likely JHS’ was 3.52 (95%CI 1.67 – 7.40, p=0.001). Given the low levels of likely JHS in males (n=3), not powered to undertake separate samples in males. This result was driven by a significant effect in females (OR 3.76, 95%CI 1.76-8.00, p=0.001).

Across the whole group ‘likely JHS’ was significantly associated with higher CIS-R (mean, sd) scores in the ‘likely JHS’ group (11.36, 9.91) compared to those without ‘likely JHS’ (5.51, 6.36) (Beta=0.091, t(3554)=5.46, p=<0.001). In analyses corrected for sex, this association remained (Beta 0.077, t(3553)=4.70, p=<0.001). In females ‘likely JHS’ was associated with higher CIS-R scores (11.97, 10.13) compared to those without (6.64, 6.88) (Beta 0.096, t(2064)=4.38, p=<0.001). (Figure 2)

##### Likely JHS and anxiety at age 18

Across the whole group ‘likely JHS’ was significantly associated with any anxiety disorder at 18 (OR 4.06, 95% CI 1.98 – 8.33, p=<0.001). Of those with ICD-10 anxiety disorder (n = 355) at 18, 11 (3.1%, 95% CI 1.74 – 5.46) had ‘likely JHS’; of those without anxiety disorder a significantly lower proportion (25, 0.78%) had ‘likely JHS’ (95%CI 0.53-1.15).

Adjusting for sex, the OR of having ICD-10 anxiety disorder given ‘likely JHS’ is 3.13 (95% CI 1.52-6.46, p=0.002). In Females OR for having ICD-10 anxiety disorder given ‘likely JHS’ was 3.34 (95% CI 1.60 – 6.96, p =0.001). See Figure 2 and Table 2.

Across the whole group ‘likely JHS’ was significantly associated with higher anxiety sensitivity (mean, sd; 55.53, 12.03) compared to those without ‘likely JHS’ (47.47, 11.82) (Beta = 0.068, t(3316)=3.95, p=<0.001). This association remained after adjusting for sex, (Beta = 0.056, t(3315)=3.27, p=0.001). In females, those with ‘likely JHS’ had higher anxiety sensitivity (55.94, 12.26) compared to those without (49.36, 11.74). (Figure 2)

## DISCUSSION

There is a robust literature linking both hypermobility with psychopathology in adults (e.g. ^30,55^). To our knowledge, our findings demonstrate for the first time a link between generalized joint hypermobility at age 14 yrs and subsequent depression in adolescents, showing this in males only. We also demonstrate the mediating effect of autonomic physiology (heart rate) on this relationship (which was robust to missing data). We found no association between generalized joint hypermobility alone and anxiety. This observation is striking as such an association is widely observed in adults (e.g. ^30,56^). However symptomatic joint hypermobility (presence of generalized joint hypermobility and chronic widespread pain) predicted both anxiety and depression in female adolescents and adjusted for sex. Unfortunately, the very small number of male cases of ‘likely JHS’ makes further interpretation difficult in males.

Hypermobility appears to be familial. Although no consistent gene is implicated, symptomatic hypermobility (hEDS/JHS/HSD) is considered to be an autosomal dominant trait with incomplete penetrance, variable expressivity, and influenced by sex, since hypermobility is more common in females ^20^: However, despite this female preponderance hypermobility is also associated with neurodevelopmental disorder that are more common in males including Fragile X syndrome ^57^, Attention Deficit Hyperactivity Disorder and Autism Spectrum Disorder ^31^. These conditions have increased vulnerability to clinical anxiety ^58^ and depression ^59^. Speculatively in males related mechanisms may account for the high rates of depression at age 18 yrs associated with generalized joint hypermobility aged 14 yrs. However, these observations contrast with early findings in adults of sex differences in psychiatric symptoms associated with hypermobility, in which higher anxiety scores and the presence of autonomic symptomatology are reported in hypermobile females compared to men ^60^.

Hypermobility is associated with dysautonomia ^35^, typically ‘postural tachycardia syndrome’ which itself enhances vulnerability to psychiatric symptoms ^37^. Adolescents with postural tachycardia are observed to have greater resting heart rates than controls ^61^. Our present finding of a mediating role of heart rate on the relationship between hypermobility and depression may also reflect a presence of dysautonomia/altered physiological arousal. Interestingly a Swedish cohort study demonstrated higher heart rate at baseline in males was associated with development of subsequent psychopathology, notably anxiety ^62^.

Our data suggest that generalized joint hypermobility may predict depression in males, but the additional presence of features of symptomatic hypermobility (i.e. chronic widespread pain) is required in females. The categorization and classification of joint hypermobility and symptomatic joint hypermobility remains the subject of substantial debate ^21^: Although the Beighton Scale remains the most widely used scoring system for measuring hypermobility, there are considerable questions about its use ^63-65^, particularly in children and adolescents ^66,67^ and cut-offs vary widely.

## CONCLUSIONS

Our study provides evidence for both a longitudinal association between a common variant of connective tissue, hypermobility, and common mental illness, depression, at age 18 yrs. We also observe the association between symptomatic joint hypermobility and depression and anxiety at age 18 yrs. In recent years there has been much attention in lay media given to hypermobility as a single biomedical cause of sometimes profound disability. Together our findings highlight the importance of a broad biopsychosocial approach to these complex conditions with the possibility of screening for hypermobility in adolescence as it may provide opportunities for early intervention to mitigate common psychiatric disorders.

## Data Availability

ALSPAC data is available from ALSPAC on request

http://www.bristol.ac.uk/alspac/

## ACKNOWLEDGEMENTS

We are extremely grateful to all the families who took part in this study, the midwives for their help in recruiting them, and the whole ALSPAC team, which includes interviewers, computer and laboratory technicians, clerical workers, research scientists, volunteers, managers, receptionists and nurses

## GOVERNANCE

Ethical approval for the study was obtained from the ALSPAC Ethics and Law Committee and the Local Research Ethics Committees. Informed consent for the use of data collected via questionnaires and clinics was obtained from participants following the recommendations of the ALSPAC Ethics and Law Committee at the time. Children were invited to give assent where appropriate. Study participants have the right to withdraw their consent for elements of the study or from the study entirely at any time. Full details of the ALSPAC consent procedures are available on the study website (http://www.bristol.ac.uk/alspac/researchers/research-ethics/).

## DATA SHARING

Data is available from ALSPAC on request http://www.bristol.ac.uk/alspac/

---

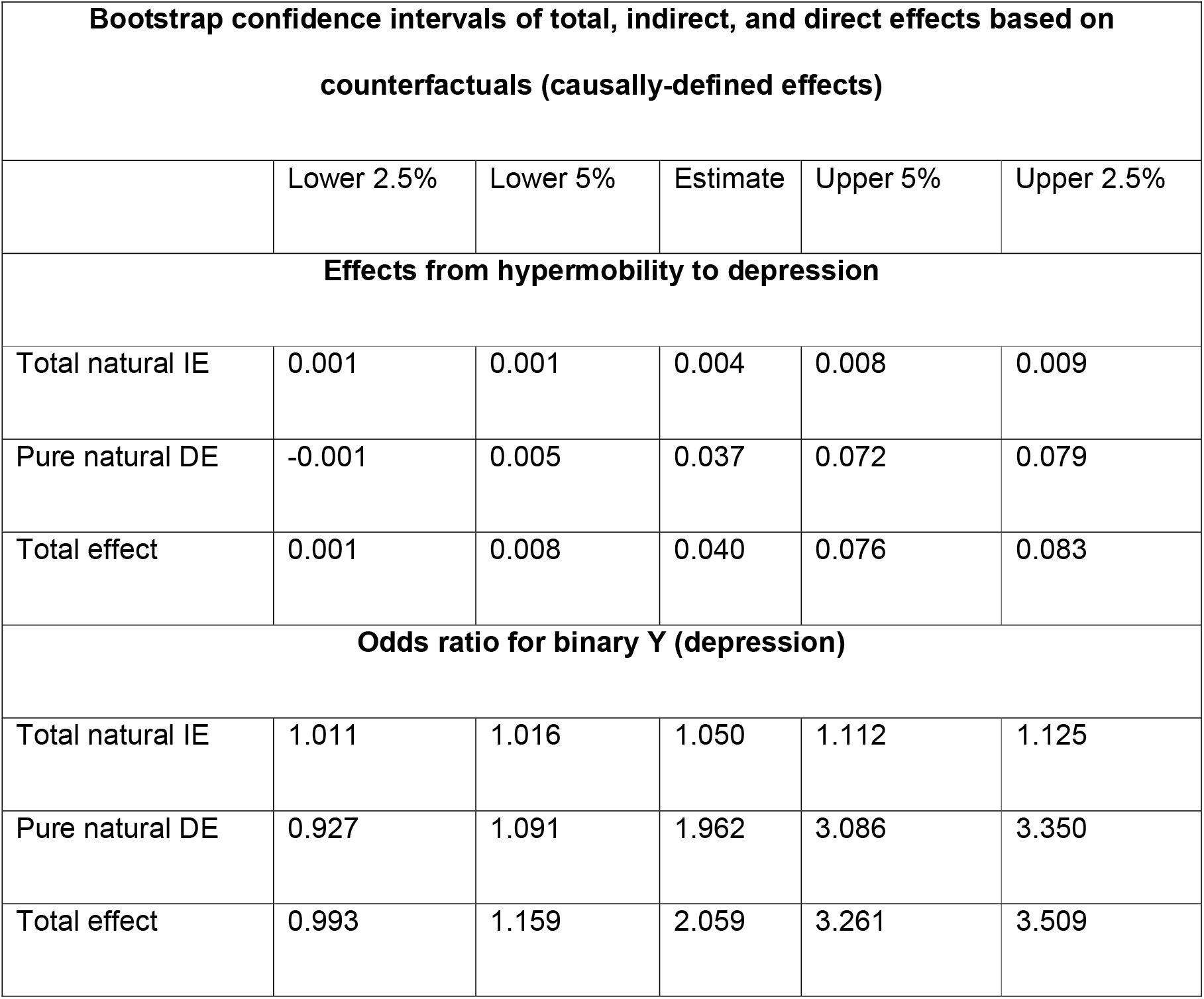

## REFERENCES

1. Murray CJ, Lopez AD. Measuring the global burden of disease. N Engl J Med. 2013;369(5):448–457.

2. Merikangas KR, He J-p, Burstein M, et al. Lifetime Prevalence of Mental Disorders in U.S. Adolescents: Results from the National Comorbidity Survey Replication–Adolescent Supplement (NCS-A). Journal of the American Academy of Child & Adolescent Psychiatry. 2010;49(10):980–989.

3. Davies SJ, Pearson RM, Stapinski L, et al. Symptoms of generalized anxiety disorder but not panic disorder at age 15 years increase the risk of depression at 18 years in the Avon Longitudinal Study of Parents and Children (ALSPAC) cohort study. Psychol Med. 2016;46(1):73–85.

4. Niarchou M, Zammit S, Lewis G. The Avon Longitudinal Study of Parents and Children (ALSPAC) birth cohort as a resource for studying psychopathology in childhood and adolescence: a summary of findings for depression and psychosis. Soc Psychiatry Psychiatr Epidemiol. 2015;50(7):1017–1027.

5. Stringaris A, Lewis G, Maughan B. Developmental pathways from childhood conduct problems to early adult depression: findings from the ALSPAC cohort. Br J Psychiatry. 2014;205(1):17–23.

6. Wiles NJ, Haase AM, Lawlor DA, Ness A, Lewis G. Physical activity and depression in adolescents: cross-sectional findings from the ALSPAC cohort. Soc Psychiatry Psychiatr Epidemiol. 2012;47(7):1023–1033.

7. Khandaker GM, Pearson RM, Zammit S, Lewis G, Jones PB. Association of serum interleukin 6 and C-reactive protein in childhood with depression and psychosis in young adult life: a population-based longitudinal study. JAMA Psychiatry. 2014;71(10):1121–1128.

8. Khandaker GM, Zammit S, Lewis G, Jones PB. Association between serum C-reactive protein and DSM-IV generalized anxiety disorder in adolescence: Findings from the ALSPAC cohort. Neurobiol Stress. 2016;4:55-61.

9. Cornish RP, John A, Boyd A, Tilling K, Macleod J. Defining adolescent common mental disorders using electronic primary care data: a comparison with outcomes measured using the CIS-R. BMJ Open. 2016;6(12):e013167.

10. Pearson RM, Evans J, Kounali D, et al. Maternal depression during pregnancy and the postnatal period: risks and possible mechanisms for offspring depression at age 18 years. JAMA Psychiatry. 2013;70(12):1312–1319.

11. McGorry PD. Is early intervention in the major psychiatric disorders justified? Yes. BMJ. 2008;337.

12. Costello EJ, Maughan B. Annual Research Review: Optimal outcomes of child and adolescent mental illness. Journal of Child Psychology and Psychiatry. 2015;56(3):324–341.

13. Grahame R. Hypermobility: an important but often neglected area within rheumatology. Nat Clin Pract Rheumatol. 2008;4(10):522–524.

14. Clinch J, Deere K, Sayers A, et al. Epidemiology of generalized joint laxity (hypermobility) in fourteen-year-old children from the UK: a population-based evaluation. Arthritis Rheum. 2011;63(9):2819–2827.

15. Kirk JA, Ansell BM, Bywaters EG. The hypermobility syndrome. Musculoskeletal complaints associated with generalized joint hypermobility. Ann Rheum Dis. 1967;26(5):419–425.

16. Beighton P, Solomon L, Soskolne CL. Articular mobility in an African population. Ann Rheum Dis. 1973;32(5):413–418.

17. Koldas Dogan S, Taner Y, Evick D. Benign joint hypermobility syndrome in patients with attention defecit/hyperactivity disorders. Turkish Journal of Rheumatology. 2011;26(3):187–192.

18. Mulvey MR, Macfarlane GJ, Beasley M, et al. Modest association of joint hypermobility with disabling and limiting musculoskeletal pain: results from a large-scale general population-based survey. Arthritis Care Res (Hoboken). 2013;65(8):1325–1333.

19. Figueroa JJ, Bott-Kitslaar DM, Mercado JA, et al. Decreased orthostatic adrenergic reactivity in non-dipping postural tachycardia syndrome. Auton Neurosci. 2014;185:107-111.

20. Castori M. Ehlers-danlos syndrome, hypermobility type: an underdiagnosed hereditary connective tissue disorder with mucocutaneous, articular, and systemic manifestations. ISRN Dermatol. 2012;2012:751768.

21. Castori M, Tinkle B, Levy H, Grahame R, Malfait F, Hakim A. A framework for the classification of joint hypermobility and related conditions. Am J Med Genet C Semin Med Genet. 2017;175(1):148–157.

22. Singer W, Shen WK, Opfer-Gehrking TL, McPhee BR, Hilz MJ, Low PA. Heart rate-dependent electrocardiogram abnormalities in patients with postural tachycardia syndrome. Auton Neurosci. 2003;103(1-2):106-113.

23. Tinkle BT, Bird HA, Grahame R, Lavallee M, Levy HP, Sillence D. The lack of clinical distinction between the hypermobility type of Ehlers-Danlos syndrome and the joint hypermobility syndrome (a.k.a. hypermobility syndrome). Am J Med Genet A. 2009;149A(11):2368-2370.

24. Grahame R, Bird HA, Child A. The revised (Brighton 1998) criteria for the diagnosis of benign joint hypermobility syndrome (BJHS). J Rheumatol. 2000;27(7):1777–1779.

25. Bulbena A, Baeza-Velasco C, Bulbena-Cabre A, et al. Psychiatric and psychological aspects in the Ehlers-Danlos syndromes. Am J Med Genet C Semin Med Genet. 2017;175(1):237–245.

26. Baeza-Velasco C, Pailhez G, Bulbena A, Baghdadli A. Joint hypermobility and the heritable disorders of connective tissue: clinical and empirical evidence of links with psychiatry. Gen Hosp Psychiatry. 2015;37(1):24–30.

27. Eccles JA, Owens AP, Mathias CJ, Umeda S, Critchley HD. Neurovisceral phenotypes in the expression of psychiatric symptoms. Front Neurosci. 2015;9:4.

28. Mallorqui-Bague N, Bulbena A, Pailhez G, Garfinkel SN, Critchley HD. Mind-Body Interactions in Anxiety and Somatic Symptoms. Harv Rev Psychiatry. 2016;24(1):53–60.

29. Bowen J, Fatjo J, Serpell JA, Bulbena-Cabre A, Leighton E, Bulbena A. First evidence for an association between joint hypermobility and excitability in a non-human species, the domestic dog. Sci Rep. 2019;9(1):8629.

30. Smith TO, Easton V, Bacon H, et al. The relationship between benign joint hypermobility syndrome and psychological distress: a systematic review and meta-analysis. Rheumatology (Oxford). 2014;53(1):114–122.

31. Cederlof M, Larsson H, Lichtenstein P, Almqvist C, Serlachius E, Ludvigsson JF. Nationwide population-based cohort study of psychiatric disorders in individuals with Ehlers-Danlos syndrome or hypermobility syndrome and their siblings. BMC Psychiatry. 2016;16:207.

32. Eccles JA, Beacher FD, Gray MA, et al. Brain structure and joint hypermobility: relevance to the expression of psychiatric symptoms. Br J Psychiatry. 2012;200(6):508–509.

33. Mallorqui-Bague N, Garfinkel SN, Engels M, et al. Neuroimaging and psychophysiological investigation of the link between anxiety, enhanced affective reactivity and interoception in people with joint hypermobility. Front Psychol. 2014;5:1162.

34. Eccles J, Owens A, Harrison N, Grahame R, Critchley H. Joint hypermobility and autonomic hyperactivity: an autonomic and functional neuroimaging study. The Lancet. 2016;387, Supplement 1:S40.

35. Gazit Y, Nahir AM, Grahame R, Jacob G. Dysautonomia in the joint hypermobility syndrome. Am J Med. 2003;115(1):33–40.

36. Celletti C, Camerota F, Castori M, et al. Orthostatic Intolerance and Postural Orthostatic Tachycardia Syndrome in Joint Hypermobility Syndrome/Ehlers-Danlos Syndrome, Hypermobility Type: Neurovegetative Dysregulation or Autonomic Failure? Biomed Res Int. 2017;2017:9161865.

37. Mathias CJ, Low DA, Iodice V, Owens AP, Kirbis M, Grahame R. Postural tachycardia syndrome--current experience and concepts. Nat Rev Neurol. 2012;8(1):22–34.

38. Tobias JH, Deere K, Palmer S, Clark EM, Clinch J. Joint hypermobility is a risk factor for musculoskeletal pain during adolescence: findings of a prospective cohort study. Arthritis Rheum. 2013;65(4):1107–1115.

39. Harris MJ. ADD/ADHD and hypermobile joints. J Paediatr Child Health. 1998;34(4):400–401.

40. Shiari R, Saeidifard F, Zahed G. Evaluation of the Prevalence of Joint Laxity in Children with Attention Deficit/Hyperactivity Disorder. Ann Paediatr Rheum. 2013;2(2):78–80.

41. Tantam D, Evered C, Hersov L. Asperger’s syndrome and ligamentous laxity. Journal of the American Academy of Child and Adolescent Psychiatry. 1990;29(6):892–896.

42. Celletti C, Mari G, Ghibellini G, Celli M, Castori M, Camerota F. Phenotypic variability in developmental coordination disorder: Clustering of generalized joint hypermobility with attention deficit/hyperactivity disorder, atypical swallowing and narrative difficulties. Am J Med Genet C Semin Med Genet. 2015;169C(1):117-122.

43. Kirby A, Davies R. Developmental Coordination Disorder and Joint Hypermobility Syndrome--overlapping disorders? Implications for research and clinical practice. Child Care Health Dev. 2007;33(5):513–519.

44. Boyd A, Golding J, Macleod J, et al. Cohort Profile: the ‘children of the 90s’--the index offspring of the Avon Longitudinal Study of Parents and Children. Int J Epidemiol. 2013;42(1):111–127.

45. Fraser A, Macdonald-Wallis C, Tilling K, et al. Cohort Profile: the Avon Longitudinal Study of Parents and Children: ALSPAC mothers cohort. Int J Epidemiol. 2013;42(1):97–110.

46. Subramanyam V, Janaki K. Joint hypermobility in south Indian children. Indian pediatrics. 1996;33:771-771.

47. Seckin U, Tur BS, Yilmaz O, Yagci I, Bodur H, Arasil T. The prevalence of joint hypermobility among high school students. Rheumatol Int. 2005;25(4):260–263.

48. Gyldenkerne B, Iversen K, Roegind H, Fastrup D, Hall K, Remvig L. Prevalence of general hypermobility in 12-13-year-old school children and impact of an intervention against injury and pain incidence. Advances in Physiotherapy. 2007;9(1):10–15.

49. Deere KC, Clinch J, Holliday K, et al. Obesity is a risk factor for musculoskeletal pain in adolescents: findings from a population-based cohort. Pain. 2012;153(9):1932–1938.

50. Lewis G, Pelosi AJ, Araya R, Dunn G. Measuring psychiatric disorder in the community: a standardized assessment for use by lay interviewers. Psychol Med. 1992;22(2):465–486.

51. Lewis G, Araya R. Classification, disability and the public health agenda. Br Med Bull. 2001;57:3-15.

52. Brugha TS, Bebbington PE, Jenkins R, et al. Cross validation of a general population survey diagnostic interview: a comparison of CIS-R with SCAN ICD-10 diagnostic categories. Psychol Med. 1999;29(5):1029–1042.

53. Muthén L, Muthén B. Statistical Analysis with Latent Variables: Mplus User’s Guide. In: Muthén & Muthén, Los Angeles, CA; 2012.

54. Muthen B, Asparouhov T. Causal Effects in Mediation Modeling: An Introduction With Applications to Latent Variables. Struct Equ Modeling. 2015;22(1):12–23.

55. Sanches SH, Osorio Fde L, Udina M, Martin-Santos R, Crippa JA. Anxiety and joint hypermobility association: a systematic review. Rev Bras Psiquiatr. 2012;34 Suppl 1:S53-60.

56. Bulbena A, Gago J, Pailhez G, Sperry L, Fullana MA, Vilarroya O. Joint hypermobility syndrome is a risk factor trait for anxiety disorders: a 15-year follow-up cohort study. Gen Hosp Psychiatry. 2011;33(4):363–370.

57. Garber KB, Visootsak J, Warren ST. Fragile X syndrome. Eur J Hum Genet. 2008;16(6):666–672.

58. van Steensel FJ, Bogels SM, Perrin S. Anxiety disorders in children and adolescents with autistic spectrum disorders: a meta-analysis. Clin Child Fam Psychol Rev. 2011;14(3):302–317.

59. Magnuson KM, Constantino JN. Characterization of depression in children with autism spectrum disorders. J Dev Behav Pediatr. 2011;32(4):332–340.

60. Sanches SB, Osorio FL, Louzada-Junior P, Moraes D, Crippa JA, Martin-Santos R. Association between joint hypermobility and anxiety in Brazilian university students: Gender-related differences. J Psychosom Res. 2014.

61. Singer W, Sletten DM, Opfer-Gehrking TL, Brands CK, Fischer PR, Low PA. Postural tachycardia in children and adolescents: what is abnormal? J Pediatr. 2012;160(2):222–226.

62. Latvala A, Kuja-Halkola R, Ruck C, et al. Association of Resting Heart Rate and Blood Pressure in Late Adolescence With Subsequent Mental Disorders: A Longitudinal Population Study of More Than 1 Million Men in Sweden. JAMA Psychiatry. 2016;73(12):1268–1275.

63. Juul-Kristensen B, Rogind H, Jensen DV, Remvig L. Inter-examiner reproducibility of tests and criteria for generalized joint hypermobility and benign joint hypermobility syndrome. Rheumatology (Oxford). 2007;46(12):1835–1841.

64. Remvig L, Engelbert RH, Berglund B, et al. Need for a consensus on the methods by which to measure joint mobility and the definition of norms for hypermobility that reflect age, gender and ethnic-dependent variation: is revision of criteria for joint hypermobility syndrome and Ehlers-Danlos syndrome hypermobility type indicated? Rheumatology (Oxford). 2011;50(6):1169–1171.

65. Remvig L, Jensen DV, Ward RC. Are diagnostic criteria for general joint hypermobility and benign joint hypermobility syndrome based on reproducible and valid tests? A review of the literature. J Rheumatol. 2007;34(4):798–803.

66. Singh H, McKay M, Baldwin J, et al. Beighton scores and cut-offs across the lifespan: cross-sectional study of an Australian population. Rheumatology (Oxford). 2017.

67. Simmonds J. Generalized joint hypermobility: a timely population study and proposal for Beighton cut-offs: Beighton cut-offs for generalized joint hypermobility. Rheumatology (Oxford). 2017.

